# Impact of Atrial Fibrillation on Hospital Outcomes in NSTEMI Patients: A Retrospective Cohort Study

**DOI:** 10.1101/2025.07.07.25331016

**Authors:** Swatam Jain, Pinak Shah, Kartika Shetty

**Affiliations:** Department of Internal Medicine, HCA Healthcare; MountainView Hospital, Las Vegas, NV, USA

**Keywords:** non-ST elevation Myocardial infarction (NSTEMI), Atrial fibrillation, Coronary artery bypass grafting (CABG), female sex

## Abstract

Atrial fibrillation (AF) is the most common sustained arrhythmia and frequently occurs in patients with acute coronary syndromes (ACS). Non-ST elevation myocardial infarction (NSTEMI) accounts for nearly 70% of ACS hospitalizations and poses significant morbidity and healthcare burden. Despite its prevalence, the impact of AF on in-hospital outcomes in NSTEMI remains underrecognized, and current risk models often exclude AF.

**Methods:** We conducted a retrospective cohort study using de-identified patient-level data from HCA Healthcare, capturing 31,649 NSTEMI admissions across 180 U.S. hospitals (2021–2022). Patients were stratified based on coronary artery bypass grafting (CABG) status. Multivariable logistic and linear regression models evaluated associations between AF and in-hospital mortality, 30-day readmission, and length of stay (LOS).

**Results:** AF was independently associated with worse in-hospital outcomes in both CABG and non-CABG groups. Among CABG patients, AF was linked to increased odds of in-hospital mortality (OR 2.02), 30-day readmission (OR 1.15), and prolonged LOS (OR 1.21). In non-CABG patients, AF was similarly associated with higher odds of mortality (OR 1.89), readmission (OR 1.23), and LOS (OR 1.31) (all p<0.05). Female sex, heart failure, CKD, and COPD were also linked to adverse outcomes.

**Conclusion:** In this large, multicenter cohort, AF was significantly associated with increased in-hospital mortality, readmission, and LOS among NSTEMI patients, irrespective of CABG status. These findings highlight AF as a key clinical factor warranting consideration in NSTEMI management. Future studies should explore mechanisms underlying these associations and identify strategies for risk mitigation in this high-risk population.

## Introduction

Non-ST elevation myocardial infarction (NSTEMI) accounts for the majority of acute coronary syndrome (ACS) hospitalizations in the United States and remains a significant contributor to morbidity, mortality, and healthcare resource utilization. Despite advancements in guideline-directed management and invasive strategies, outcomes such as hospital readmission, in-hospital mortality, and prolonged length of stay remain areas of concern. Atrial fibrillation (AF), the most common sustained arrhythmia worldwide, frequently coexists with coronary artery disease (CAD) and ACS.

While atrial fibrillation (AF) and coronary artery disease (CAD) share several common risk factors—including hypertension, diabetes, and aging—the nature of their relationship remains incompletely understood. [1,2] AF has been identified as a potential risk factor for the development of CAD, likely mediated by systemic inflammation, endothelial dysfunction, and a prothrombotic state. However, the role of CAD as a precursor to AF has yielded inconsistent findings across studies. [3-5] Prior investigations have shown that patients with both AF and CAD often exhibit more advanced atherosclerotic burden and face worse clinical outcomes, although the precise magnitude of this impact has not been consistently defined. [4,6] AF is present in approximately 6–21% of patients hospitalized with acute myocardial infarction, and its presence has been linked to increase in-hospital mortality, prolonged length of stay (LOS), and greater healthcare utilization. [7]

Existing clinical tools such as the TIMI and GRACE risk scores—used widely for risk stratification in NSTEMI—do not include AF as a risk-enhancing variable, potentially overlooking patients with higher short-term risk. Moreover, prior research has often examined broad ACS populations or focused on long-term outcomes without adequately characterizing short-term in-hospital trajectories in NSTEMI patients with AF.

This study addresses a critical knowledge gap by evaluating the influence of atrial fibrillation on early hospital outcomes in NSTEMI patients within a contemporary clinical context. We hypothesized that atrial fibrillation would be independently associated with adverse early hospital outcomes—including increased in-hospital mortality, longer length of stay, and higher 30-day readmission rates—among patients admitted with NSTEMI. By clarifying the prognostic value of AF in this high-risk population, the study lays a foundation for refining risk stratification tools, enhancing individualized care pathways, and guiding resource allocation. These findings may prompt renewed attention to AF as a modifiable variable in acute cardiac care and inform future updates to clinical practice guidelines aimed at improving outcomes in NSTEMI.

## Methods

### Data source

A retrospective analysis was conducted from January 1, 2021 – December 31, 2022, using de-identified data from the nationwide HCA Healthcare in the United States. Data was collected from 180 hospitals within HCA Healthcare enterprise in the United States. Protocol was submitted to IRB and the research activity was determined to exempt from Institutional Review Board oversight in accordance with institutional regulations and policy.

### Study Patients

Inclusion criteria for this study was age over 18, admission diagnosis of NSTEMI and had received coronary angiography during index admission. This was done using the International Classification of Disease□ICD-10 Clinical Modification CM codes. Exclusion criteria for this study was a diagnosis of STEMI during index admission (see supplemental table 1 & 2 for ICD-10 codes).

**Table 1.**
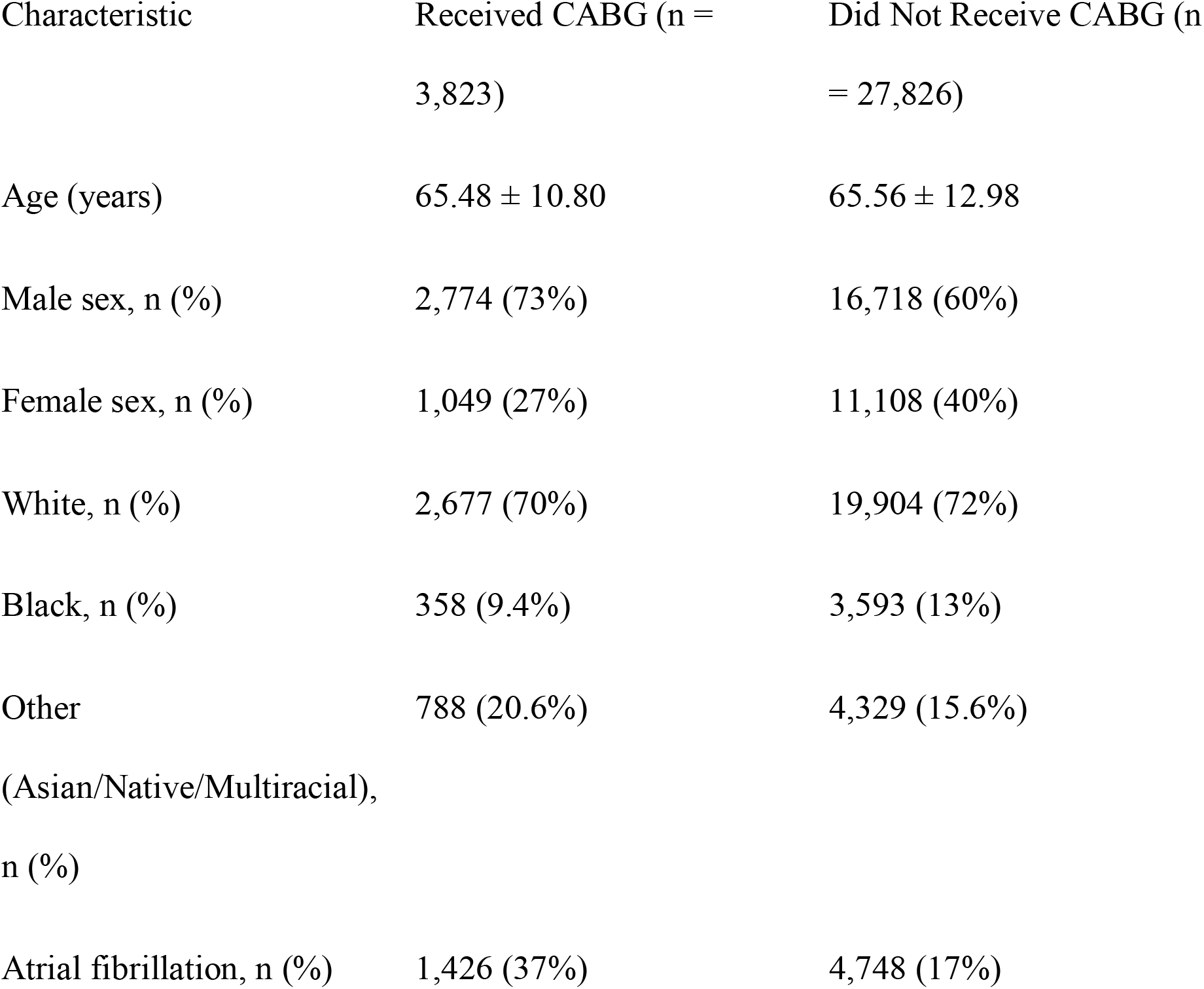
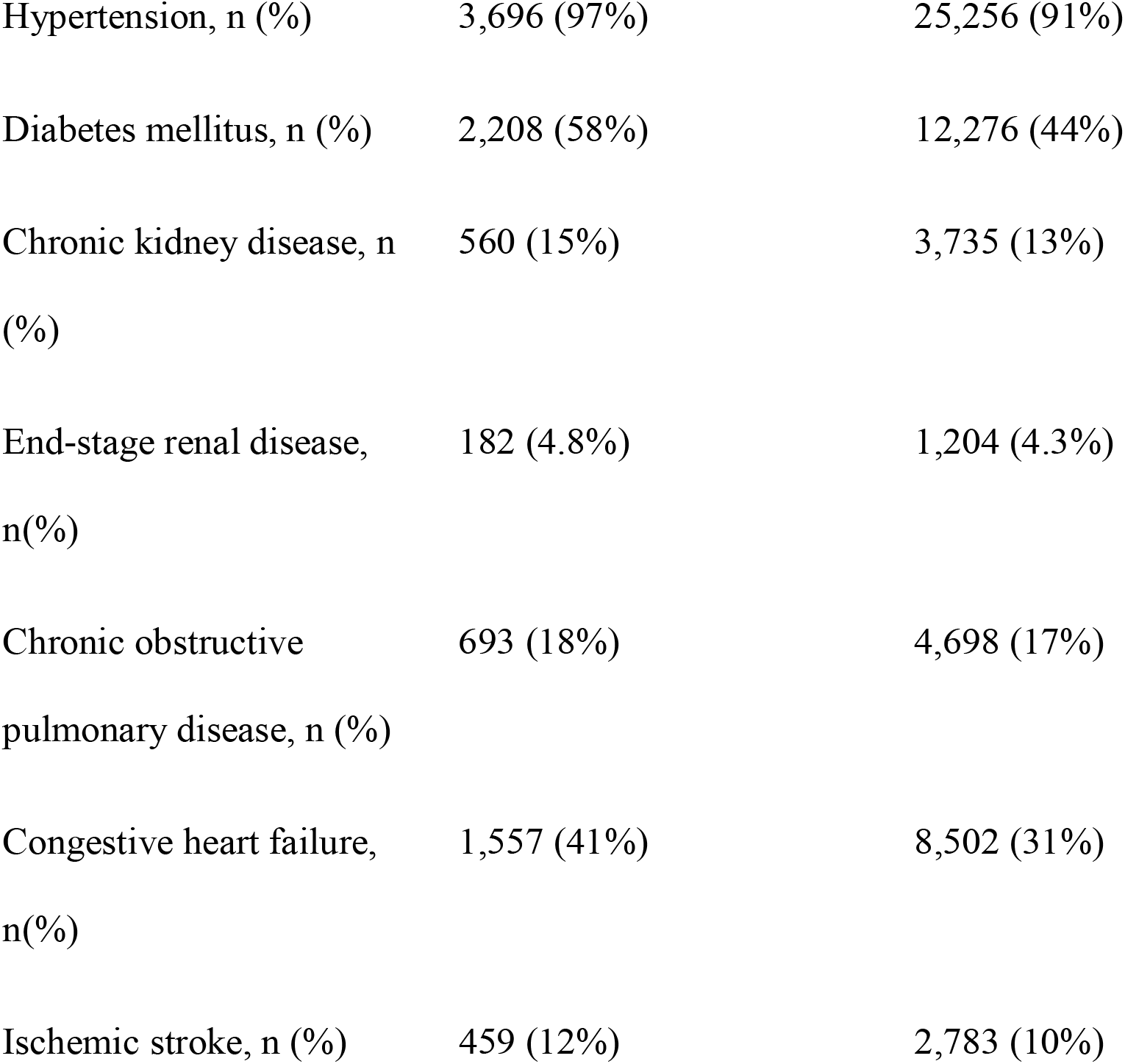
Baseline Characteristics of NSTEMI Patients by CABG Status.

| Characteristic | Received CABG (n =<br>3,823) | Did Not Receive CABG (n<br>= 27,826) |
| --- | --- | --- |
| Age (years) | 65.48 ± 10.80 | 65.56 ± 12.98 |
| Male sex, n (%) | 2,774 (73%) | 16,718 (60%) |
| Female sex, n (%) | 1,049 (27%) | 11,108 (40%) |
| White, n (%) | 2,677 (70%) | 19,904 (72%) |
| Black, n (%) | 358 (9.4%) | 3,593 (13%) |
| Other<br>(Asian/Native/Multiracial),<br>n (%) | 788 (20.6%) | 4,329 (15.6%) |
| Atrial fibrillation, n (%) | 1,426 (37%) | 4,748 (17%) |
| Hypertension, n (%) | 3,696 (97%) | 25,256 (91%) |
| Diabetes mellitus, n (%) | 2,208 (58%) | 12,276 (44%) |
| Chronic kidney disease, n (%) | 560 (15%) | 3,735 (13%) |
| End-stage renal disease, n(%) | 182 (4.8%) | 1,204 (4.3%) |
| Chronic obstructive pulmonary disease, n (%) | 693 (18%) | 4,698 (17%) |
| Congestive heart failure, n(%) | 1,557 (41%) | 8,502 (31%) |
| Ischemic stroke, n (%) | 459 (12%) | 2,783 (10%) |

**Table 2:** Hospital outcomes of patients with Atrial fibrillation vs without Atrial Fibrillation.

|  | Received CABG |  | Didn't Receive CABG |  |
| --- | --- | --- | --- | --- |
|  | Odds Ratio | p-Value | Odds Ratio | p-Value |
| Mortality | 2.02 (95% CI, 1.43-2.87) | <0.001 | 1.89 (95% CI, 1.59-2.23) | <0.001 |
| Readmission | 1.15 (95% CI, 1.01-1.32) | 0.031 | 1.23 (95% CI, 1.15-1.32) | <0.001 |
| Length of Stay | 1.21 (95% CI, 1.18-1.25) | <0.001 | 1.31 (95% CI, 1.28-1.34) | <0.001 |

### Study Design

This investigation was structured as a retrospective cohort study to evaluate the impact of atrial fibrillation on key clinical outcomes among NSTEMI patients who either received coronary artery bypass grafting (CABG) or did not receive CABG. We stratified the NSTEMI cohort into two mutually exclusive groups—those who underwent CABG during the index admission (CABG cohort) and those who did not (non-CABG cohort).

### Study Cohorts

Cohort 1 included 3,823 NSTEMI patients who underwent CABG during the index hospitalization, while cohort 2 included 27,826 NSTEMI patients who did not undergo CABG. All patients in this analysis had a primary diagnosis of NSTEMI. After using multivariable adjustment based on age, race, hypertension, diabetes mellitus, chronic kidney disease, chronic obstructive pulmonary disease, and end-stage renal disease, each cohort included 3,823 patients.

Baseline demographics (age, race, comorbidities) were summarized for each cohort (see Supplementary Table 2).

### Predictor and Outcome Variables

The primary predictor in this study was the presence of atrial fibrillation (AF) during the index hospitalization, identified using ICD-10-CM codes. The primary outcomes included in-hospital mortality (dichotomized as yes/no), 30-day hospital readmission (yes/no), and length of stay (LOS), recorded as a continuous variable in days. Additional covariates adjusted for in the analysis included age, sex, race, hypertension, diabetes mellitus, chronic kidney disease, ischemic stroke, chronic obstructive pulmonary disease, and end-stage renal disease, all treated as binary variables.

### Statistical Analysis

To reduce bias and ensure balanced comparison between the CABG and non-CABG cohorts, we employed multivariable regression adjustment using variables including age, race, hypertension, diabetes mellitus, chronic kidney disease, chronic obstructive pulmonary disease, and end-stage renal disease. This created comparable groups, enabling a focused analysis on the impact of atrial fibrillation on hospital outcomes.

Length of stay was analyzed via log-gamma regression in the CABG cohort and negative binomial regression in the non-CABG cohort. In-hospital mortality and 30-day readmission were assessed using multivariable logistic regression, adjusting for age, race, hypertension, diabetes mellitus, chronic kidney disease, chronic obstructive pulmonary disease, and end-stage renal disease. Continuous outcomes are presented as multiplicative effects or incidence rate ratios with 95% confidence intervals; binary outcomes are reported as odds ratios with 95% confidence intervals. Statistical significance was defined as p < 0.05.

## Results

The total number of patients included in the study were 31,649. There were 3,823 patients who received CABG and 27,826 patients who did not receive CABG (see table 2).

### CABG group

For patients who received CABG, 37% of patients had atrial fibrillation and atrial fibrillation was a significant predictor of mortality, 30-Day readmission and length of stay. Patients with atrial fibrillation were 2.02 (95% CI [1.43-2.87], p<0.001) times more likely to experience mortality compared to without atrial fibrillation (see table 2). The odds of patients with atrial fibrillation being readmitted within 30 days was 1.15(95% CI [1.01-1.32], p=0.031) times higher than patients without atrial fibrillation. The odds of patients with atrial fibrillation for days spent in the hospital was 1.21(95% CI [1.18-1.25], p<0.001) times higher patients without atrial fibrillation.

### Non CABG group

For patients who didn’t receive CABG, 17% of patients had atrial fibrillation and atrial fibrillation was a significant predictor of mortality, 30-Day readmission and length of stay. The odds of patients with atrial fibrillation experiencing mortality were 1.89 (95% CI [1.59-2.23], p<0.001) times higher patients without atrial fibrillation. The odds of patients with atrial fibrillation being readmitted within 30 days was 1.23(95% CI [1.15-1.32], p<0.001) times higher than patients without atrial fibrillation. The odds of patients with atrial fibrillation for days spent in the hospital was 1.3131 (95% CI [1.28-1.34], p<0.001) times higher than patients without atrial fibrillation.

For patients who received CABG, 27% of patients were female and female sex was a significant predictor of mortality and Length of stay. The odds of female patients experiencing mortality was 1.66(95% CI [1.165-2.362], p<0.005) times higher than male patients (see table 3). The odds of female patients for days spent in the hospital was 1.064(95% CI [1.030-1.099], p<0.001) times higher than male patients. For patients who didn’t receive CABG, 40% of patients were female and female sex was a significant predictor of 30-Day readmission and Length of stay. The odds of female patients being readmitted within 30 days was 1.19(95% CI [1.129-1.263], p<0.001) times higher male patients. The odds of female patients for days spent in the hospital was 1.075(95% CI [1.056-1.95], p<0.001) times higher than male patients (see Table 3).

**Table 3:** Hospital outcomes of Length of stay, Readmission mortality with other co-variates.

| Hospital Outcomes Intercept | Received CABG |  |  | Didn't Receive CABG |  |  |
| --- | --- | --- | --- | --- | --- | --- |
|  | Length of Stay | Readmission | Mortality | Length of Stay | Readmission | Mortality |
| Female vs Male | 1.064 (95% CI, [1.030-1.099], $p<0.001$) | 1.110 (95% CI, [0.961-1.281], $p=0.156$) | 1.666 (95% CI [1.165-2.362], $p<0.005$) | 1.075 (95% CI, [1.056-1.95], $p<0.001$) | 1.194 (95% CI, [1.129-1.263], $p<0.001$) | 1.016 (95% CI [0.864-1.193], $p=0.845$) |
| Diabetes Mellitus: Diagnosed (vs. Not) | 1.050 (95% CI [1.019-1.083], $p=0.001$) | 1.106 (95% CI [0.968-1.265], $p=0.138$) | 0.948 (95% CI [0.671-1.394], $p=0.766$) | 1.124 (95% CI [1.103-1.145], $p<0.001$) | 1.078 (95% CI [1.018-1.142], $p=0.001$) | 1.236 (95% CI [1.046-1.464], $p=0.013$) |
| Hypertension: Diagnosed (vs. Not) | 1.077 (95% CI [0.992-1.167], $p=0.075$) | 1.042 (95% CI [0.728-1.498], $p=0.822$) | NA | 1.122 (95% CI [1.064-1.160], $p<0.001$) | 1.195 (95% CI [1.077-1.329], $p=0.001$) | 1.665 (95% CI [1.093-2.683], $p=0.025$) |
| Congestive Heart Failure: Diagnosed (vs. Not) | 1.190 (95% CI [1.154-1.227], $p<0.001$) | 0.925 (95% CI [0.807-1.055], $p=0.238$) | 1.937 (95% CI [1.365-2.769], $p<0.001$) | 1.634 (95% CI [1.603-1.666], $p<0.001$) | 1.354 (95% CI [1.274-1.440], $p<0.001$) | 3.482 (95% CI [2.919-4.166], $p<0.001$) |
| Ischemic Stroke: Diagnosed (vs. Not) | 1.577 (95% CI [1.438-1.734], $p<0.001$) | 1.112 (95% CI [0.736-1.681], $p=0.613$) | NA | 2.407 (95% CI [2.255-2.597], $p<0.001$) | 1.313 (95% CI [1.002-1.707], $p=0.045$) | 4.159 (95% CI [2.697-6.198], $p<0.001$) |
| Chronic Obstructive Pulmonary Disease: Diagnosed | 1.141 (95% CI [1.098-1.185], $p<0.001$) | 1.130 (95% CI [0.957-1.334], $p=0.151$) | 2.066 (95% CI [1.428-2.955], $p<0.001$) | 1.250 (95% CI [1.222-1.279], $p<0.001$) | 1.220 (95% CI [1.136-1.310], $p<0.001$) | 1.571 (95% CI [1.318-1.868], $p<0.001$) |
| (vs. Not) |  |  |  |  |  |  |
| Chronic<br>Kidney<br>Disease:<br>Diagnosed<br>(vs. Not) | 1.192 (95%<br>CI [1.143-1.243],<br>p<0.001) | 1.121 (95% CI<br>[0.932-1.347],<br>p=0.225) | NA | 1.342 (95% CI<br>[1.309-1.375],<br>p<0.001) | 1.269 (95% CI<br>[1.172-1.373],<br>p<0.001) | 1.708 (95%<br>CI [1.410-2.060],<br>p<0.001) |
| End-Stage<br>Renal<br>Disease:<br>Diagnosed<br>(vs. Not) | 1.587 (95%<br>CI [1.482-1.702],<br>p<0.001) | 0.991 (95% CI<br>[0.730-1.343],<br>p=0.953) | NA | 1.701 (95% CI<br>[1.637-1.765],<br>p<0.001) | 1.594 (95% CI<br>[1.405-1.806],<br>p<0.001) | 2.194 (95%<br>CI [1.678-2.837],<br>p<0.001) |

Several clinical comorbidities, including diabetes mellitus, congestive heart failure, chronic obstructive pulmonary disease, chronic kidney disease, and end-stage renal disease, were associated with prolonged hospital stays. Factors such as hypertension and diabetes mellitus were found to be significant predictors of experiencing mortality (see Table 3).

## Discussion

This study demonstrates that atrial fibrillation (AF) is significantly associated with adverse hospital outcomes—including higher in-hospital mortality, increased 30-day readmission, and longer length of stay—among patients admitted with non-ST elevation myocardial infarction (NSTEMI), regardless of whether they underwent coronary artery bypass grafting (CABG). These findings underscore the prognostic significance of AF in acute coronary syndromes (ACS) and support its consideration as a risk-modifying factor in NSTEMI management.

AF is the most common sustained arrhythmia, with an estimated prevalence of 3–6 million in the United States—a number expected to rise to over 16 million by 2050. [8] It is frequently encountered in the setting of ACS, with studies reporting a prevalence of 6–21% in acute myocardial infarction. [7] AF and coronary artery disease (CAD) share multiple risk factors— including hypertension, diabetes mellitus, obesity, and aging—but their relationship appears bidirectional. While AF has been identified as an independent risk factor for incident CAD through mechanisms such as systemic inflammation, endothelial dysfunction, and platelet activation [3,9-11], CAD and myocardial ischemia may also precipitate AF via atrial remodeling and autonomic dysregulation. [1,12] Regardless of causality, the presence of AF in CAD patients has consistently been associated with more severe coronary disease, higher SYNTAX scores, and worse clinical outcomes. [4,6]

Several inflammatory and thrombotic pathways have been implicated in linking AF to myocardial infarction (MI). Elevated levels of C-reactive protein, interleukins, and tumor necrosis factor-alpha contribute to a pro-inflammatory milieu that promotes atherosclerosis and thrombogenesis. Concurrently, markers of platelet activation—such as soluble CD40L and P-selectin—have been associated with both AF and acute coronary events. [3,9-11] These mechanisms may underlie AF’s role in precipitating type 1 or type 2 MI, whether through coronary embolism, demand ischemia, or accelerated plaque instability. [13,14]

By stratifying patients by CABG status and controlling for multiple comorbidities, our study adds real-world granularity to AF’s impact on NSTEMI outcomes, including readmissions and length of stay. Crucially, this demonstrates that AF increases adverse outcomes irrespective of CABG surgery, underscoring the arrhythmia’s independent prognostic importance. Notably, AF occurs in 5–40% of patients after CABG and is independently associated with worse outcomes in that population as well. [15,16]

In addition to AF, female sex emerged as an independent predictor of increased mortality and longer hospitalization, particularly among patients undergoing CABG. This is consistent with national data showing worse short-term outcomes among women after cardiac surgery, which may be due to delays in recognition, lower revascularization rates, and higher operative risk profiles. [17,18] Our findings underscore the need to address persistent sex-based disparities in cardiovascular care.

Despite their widespread use for short- and medium-term risk prediction in acute coronary syndromes (TIMI: ≈14-day risk of death or recurrent MI; HEART: 6-week MACE in chest-pain patients; GRACE: in-hospital to 3-year mortality post-ACS), these models remain limited in predictive specificity (36–56%) and none include atrial fibrillation as a variable. [19,20] Given AF’s demonstrated influence on mortality, LOS, and readmission, its integration into future predictive models may enhance risk discrimination, especially in intermediate-risk NSTEMI populations.

Beyond its clinical implications, AF in NSTEMI imposes a substantial economic burden. Index hospitalizations for NSTEMI average over $18,000, with prolonged LOS and early readmissions significantly driving cost. [21] Up to 15% of NSTEMI patients are readmitted within 30 days, despite advances in revascularization and medical therapy. [22,23] Targeting high-risk subgroups—such as those with AF—through enhanced rhythm monitoring, optimized anticoagulation, and tailored discharge planning may reduce unnecessary utilization and improve outcomes.

From a systems perspective, reducing readmissions and LOS is not only clinically important but financially imperative. Readmission results in loss of patient trust, unnecessary costs, and strains limited healthcare resources. [24,25] Identifying AF as a key contributor to these outcomes opens the door to targeted interventions—including remote monitoring, early follow-up, and structured rhythm management pathways.

Finally, the interplay between AF and CAD may represent a vicious cycle, where each condition exacerbates the other and amplifies adverse outcomes. [12] Disrupting this cycle through proactive management of both entities—rather than in silos—may be critical to improving patient care.

### Study strengths and Limitations

The primary strength of this study lies in its large, nationally representative cohort of 31,649 NSTEMI hospitalizations from a multihospital dataset across the United States. This extensive sample size enhances generalizability and permits robust subgroup analyses, including CABG stratification. To our knowledge, this is the largest retrospective cohort study specifically evaluating the impact of atrial fibrillation (AF) on in-hospital mortality, 30-day readmission, and length of stay in NSTEMI patients. Use of multivariable regression models strengthens the validity of the findings by minimizing confounding and ensuring balanced comparison groups.

However, limitations include the inability to distinguish new-onset from pre-existing AF, as well as the lack of data on anticoagulation use, rhythm control strategies, and AF timing relative to NSTEMI presentation. Additionally, the retrospective design and reliance on administrative data may introduce misclassification, residual confounding, and coding inaccuracies, limiting causal inference.

## Conclusion

In this large retrospective cohort analysis of patients admitted with NSTEMI, atrial fibrillation was independently associated with increased in-hospital mortality, higher 30-day readmission rates, and prolonged length of stay—regardless of whether patients underwent CABG. These findings highlight AF as a clinically significant predictor of adverse outcomes in NSTEMI and support its inclusion in future risk stratification models. Integrating AF into clinical decision-making may enhance individualized care and inform strategies to reduce readmissions and optimize resource utilization.

## Data Availability

All data produced in the present study are available upon reasonable request to the authors

## Abbreviation Full Form

ACS: Acute Coronary Syndrome
CABG: Coronary Artery Bypass Grafting
CAD: Coronary Artery Disease
CKD: Chronic Kidney Disease
COPD: Chronic Obstructive Pulmonary Disease
GRACE: Global Registry of Acute Coronary Events
LOS: Length of Stay
NSTEMI: Non-ST-Elevation Myocardial Infarction
PCI: Percutaneous Coronary Intervention
STEMI: ST-Elevation Myocardial Infarction
TIMI: Thrombolysis in Myocardial Infarction

## Supplementary Material

**Supplement Table 1:** ICD-10 Diagnosis codes

|  |  |
| --- | --- |
| ATRIAL FIBRILLATION | I48 |
| DIABETES MELLITUS | E08-E13 |
| CONGESTIVE HEART FAILURE | I150 |
| ISCHEMIC STROKE | I63 |
| HYPERTENSION | I10-I16 |
| COPD | J41,J42,J43,J44 |
| CKD | N18 (EXCLUDE N18.6) |
| ESRD | N18.6 |

**Supplement Table 2:** ICD-10 Procedure codes

|  |  |
| --- | --- |
| CORONARY ARTERY<br>BYPASS GRAFT | 33533, 33534, 33535, 33536 33510, 33511, 33512, 33513,<br>33514, 33516, 33517, 33518, 33519, 33521, 33522, 33523 |

**Supplement Table 3:** Hospital outcomes of patients receiving CABG versus patients who did not receive CABG

| Patient Characteristic | Received CABG |  | Didn't Receive CABG |  |
| --- | --- | --- | --- | --- |
|  | Overall, N =<br>3,823<sup>1</sup> | p-Value<sup>2</sup> | Overall, N =<br>27,826<sup>1</sup> | p-Value<sup>2</sup> |
| Patient Mortality, n(%) | 141 (3.7%) | 0.91 | 654 (2.4%) | 0.41 |
| Total Length of Stay in Days, Median (QR) | 11.00 (8.00 - 15.00) | <0.001 | 2.00 (2.00 - 4.00) | <0.001 |
| All-Cause Readmission Within 30 Days, n(%) | 1,801 (47%) | 0.14 | 6,960(25%) | 0.12 |

## Acknowledgments

The authors declare no acknowledgments.

